# Exploring Cultural and Linguistic Equivalence of a Translated Birth Preparedness and Complication Readiness Education Manual among Hehe Pregnant Women in Rural Tanzania: A Qualitative Study

**DOI:** 10.64898/2026.08.31.26361858

**Authors:** Rosalia Batista Mwenda, Saada Ali Seif, Rehema Ogha Stephano, Fabiola Vincent Moshi

**Author notes:** Corresponding author: Rosalia Batista Mwenda. Authors’ Email Addresses: Saada Ali Seif, Rehema Ogha Stephano, Fabiola Vincent Moshi.

## Abstract

**Background:** Birth Preparedness and Complication Readiness (BPCR) education is an important component of antenatal care. However, health education materials translated from one language into another may lose their intended meaning if linguistic, cultural, experiential, and sociolinguistic differences are not considered. In Tanzania, maternal health education is commonly delivered in Swahili, while many source materials are developed in English. This study explored the cultural and linguistic equivalence of BPCR terminology and concepts in a translated Swahili BPCR education manual among Hehe pregnant women in the rural Iringa Region, Tanzania.

**Methods:** A descriptive qualitative study was conducted in seven villages across Kilolo and Mufindi districts of Iringa Region. Seven focus group discussions (FGDs) involving 56 pregnant women were conducted. Participants were purposively selected from the Hehe community and were asked to interpret terminology and concepts contained in a harmonized Swahili BPCR education manual. The translation and adaptation process comprised six sequential steps: forward translation, synthesis, back translation, expert review, community exploration, and finalization. FGDs were conducted in Swahili by trained facilitators fluent in both Swahili and Hehe, audio-recorded with consent, transcribed, and thematically analyzed using Braun and Clarke’s six-phase approach. Analysis focused on semantic, conceptual, experiential, and sociolinguistic equivalence. Reporting was informed by the Consolidated Criteria for Reporting Qualitative Research (COREQ).

**Results:** Five themes were developed: (1) culturally and linguistically familiar expressions conveyed BPCR concepts; (2) experiential and contextual language shaped descriptions of danger signs; (3) sociolinguistic norms and modesty influenced communication about sensitive health topics; (4) some clinically important concepts had partial or limited conceptual equivalence; and (5) unfamiliar concepts required supplementary explanation. Participants identified culturally familiar expressions including *“matazamio ya kujifungua”* (“anticipated date of delivery”), *“fedha ndiyo usafiri”* (“money itself is transport”), *“chupa imepasuka”* (“the water bag has burst”), *“mtoto kutokucheza tumboni”* (“the baby is not moving in the womb”), and *“sehemu za siri”* (“private parts”). Some expressions were familiar but broader than their biomedical equivalents, while cord prolapse and neonatal cyanosis had no readily recognized community equivalents.

**Conclusion:** The findings indicate that cultural and linguistic equivalence cannot be achieved through literal translation alone. Community exploration identified expressions that were familiar and socially acceptable while also revealing clinical concepts requiring additional explanation. The findings informed refinement of the Swahili BPCR education manual while preserving the intended clinical meaning. The adapted terminology should subsequently be evaluated separately for its effects on knowledge, attitudes, practices, and other health outcomes.

## Background

Maternal and newborn health remains an important public health priority, particularly in settings where women and newborns may experience delays in recognizing complications and accessing appropriate care [1, 2, 3]. Antenatal care (ANC) provides an important opportunity to prepare women and families for childbirth, identify potential complications, and provide information about appropriate care and referral [4]. Birth Preparedness and Complication Readiness (BPCR) is an important component of maternal health education because it addresses preparation for childbirth, recognition of danger signs, identification of appropriate sources of care, and arrangements for accessing care when complications occur [5, 6].

BPCR education is commonly communicated through health workers, written materials, counselling sessions, and community health programs [7]. However, effective communication requires more than the transfer of information from one language to another. Health terminology may have different meanings, levels of familiarity, or social acceptability across linguistic and cultural settings (8). A literal translation may therefore be grammatically correct while remaining unfamiliar, ambiguous, or inappropriate for the population for whom the material is intended [9. 10, 11].

This issue is particularly relevant in multilingual health systems. In Tanzania, Swahili is widely used in health service delivery, while many health guidelines and educational resources originate in English [7, 12]. Consequently, health information may pass through several stages of translation and interpretation before reaching pregnant women during ANC [13, 14]. Differences between biomedical terminology and everyday language may affect how women interpret concepts relating to pregnancy, childbirth, postpartum care, and newborn danger signs [15, 16].

The challenge is not limited to vocabulary. Cultural meanings, social norms, lived experiences, and locally accepted ways of discussing sensitive topics may influence whether a translated expression adequately communicates the intended concept [17]. This is particularly relevant to maternal and reproductive health, where discussions may involve private body parts, bleeding, sexual and reproductive functions, labor, fetal movement, and newborn abnormalities [18, 19].

Iringa Region provides an important context for examining cultural and linguistic equivalence. The region is predominantly rural and is home to a large Hehe population [20]. Hehe and Swahili are used in everyday communication, particularly in rural communities [20, 21]. Pregnant women attending ANC commonly receive health education in Swahili [22]. However, formal biomedical terminology may not always correspond to the expressions women use when discussing pregnancy and childbirth in everyday life [23, 24].

Previous research has documented variation in BPCR knowledge and practices among pregnant women in Tanzania and elsewhere in sub-Saharan Africa [7, 25]. Although these studies provide important information about levels of BPCR knowledge, less attention has been given to how pregnant women themselves interpret and express the terminology used in translated BPCR education materials [26, 27].

Cultural equivalence approaches emphasize the preservation of intended meaning while adapting communication to the linguistic and cultural context of the target population [9, 28]. Nida’s theory of dynamic equivalence proposes that translated messages should communicate meaning in a way that is natural and meaningful to the receiving audience [29]. In the present study, this principle informed a structured translation and cultural adaptation process followed by community exploration of the harmonized Swahili BPCR education manual.

The study did not seek to determine whether cultural adaptation improved BPCR knowledge, attitudes, practices, health-seeking behaviors, or maternal and newborn outcomes. Instead, it examined how intended users interpreted translated terminology and whether culturally familiar expressions could communicate the intended BPCR concepts without compromising their clinical meaning.

The aim of this study was to explore the cultural and linguistic equivalence of BPCR terminology and concepts in a harmonized Swahili BPCR education manual among Hehe pregnant women in the rural Iringa Region, Tanzania, and to identify culturally meaningful expressions and concepts requiring additional contextual explanation.

## Methods

### Study design

A descriptive qualitative design was used to explore participants’ interpretations of translated BPCR terminology and concepts. Focus Group Discussions (FGDs) were selected because they allowed participants to describe, compare, and discuss terminology collectively and provided insight into socially shared linguistic and cultural practices. The study was conducted as the community-exploration component of a broader process of translating and culturally adapting a BPCR education manual for use among Hehe pregnant women.

### Study setting

The study was conducted in seven villages in Kilolo and Mufindi districts of Iringa Region, southern Tanzania. The region is predominantly rural and has a large Hehe population. Hehe and Swahili are commonly used in everyday communication [30]. The two districts are served by a network of public health facilities providing maternal and reproductive health services (31). Pregnant women attending ANC routinely receive health education concerning pregnancy, childbirth, danger signs, and preparation for delivery [7, 31]. The community exploration was conducted in the context of adapting a Swahili BPCR education manual for use among pregnant women from the Hehe community.

### Theoretical orientation

The study was guided by Nida’s Theory of Dynamic Equivalence, which emphasizes communicating the intended meaning of a source message in a form that is natural and meaningful to the target audience [29]. The translation was operationalized using the Cultural Equivalence Model by Nida [29] for translation of health materials. The framework was used to guide identification of terminology that was unfamiliar, culturally inappropriate, semantically incomplete, or insufficiently aligned with the intended clinical concept. The approaches allowed the study to distinguish between linguistic similarity and meaningful equivalence within the target cultural context.

### Participants and eligibility criteria

Participants were pregnant women from the Hehe community who were attending ANC in the study area. Women were eligible if they had lived in Iringa Region for at least six months, were willing and able to participate, and provided informed consent. The study focused primarily on women with previous pregnancy and childbirth experience because their experiences provided opportunities to identify community terminology associated with pregnancy, childbirth, postpartum care, and newborn care. Women employed within the health system were excluded because their professional familiarity with biomedical BPCR terminology could influence their interpretation of the translated material. Women who were unable or unwilling to participate were also excluded.

The inclusion and exclusion criteria were designed to ensure that participants could provide community-based rather than professional interpretations of the BPCR terminology.

### Sampling and sample size

Purposive sampling was used to recruit participants who met the eligibility criteria and could provide relevant information about community terminology and interpretations [32]. Seven FGDs were conducted in seven villages across Kilolo and Mufindi districts. A total of 56 pregnant women participated in the seven FGDs. The final number of FGDs was informed by the richness and repetition of information emerging during data collection [33, 34]. Data collection continued until the research team judged those additional discussions were generating limited additional information concerning culturally relevant terminology and concepts [33, 34]. The research assistants who facilitated the discussions were not counted as study participants.

### Translation and cultural adaptation process

The BPCR education manual was originally developed in English and was translated and culturally adapted for use among Hehe pregnant women in the rural Iringa Region. The process was guided by Nida’s Theory of Dynamic Equivalence and operationalized using the Cultural Equivalence translation model [29]. The translation and adaptation involved a multidisciplinary team with expertise in maternal and reproductive health, nursing, linguistics, health education, and translation. The process was designed to preserve the intended clinical meaning of the original English content while identifying linguistic, cultural, experiential, and sociolinguistic differences that could affect its interpretation by the target population [35]. Community exploration with Hehe’s pregnant women constituted a distinct stage of the process and was undertaken after translation, synthesis, back-translation, and expert review. The complete process comprised six sequential steps: forward translation, synthesis, back-translation, expert review, community exploration, and finalization. The complete process comprised six sequential steps: forward translation, synthesis, back-translation, expert review, community exploration, and finalization. The translation and cross-cultural adaptation procedures were adapted from the approach described by Beaton et al. [36], with community exploration incorporated to identify culturally meaningful terminology and assess its correspondence with the intended clinical concepts. The following are the six steps:

#### Step 1: Forward translation

The original English BPCR education content was independently translated into Swahili by two forward translators. The translators were selected based on their relevant language and subject-matter expertise and considered both linguistic accuracy and the appropriateness of BPCR terminology for the intended target population.

#### Step 2: Synthesis

The two forward translations were compared and synthesized into a single harmonized Swahili version. Differences between the translations were reviewed and resolved, and the decisions were documented in a translation synthesis record.

#### Step 3: Back translation

The harmonized Swahili BPCR material was independently back-translated into English by two translators who had not participated in the forward translation. The back-translations were compared with the original English content to identify discrepancies in meaning, terminology, and conceptual interpretation.

#### Step 4: Expert review

An expert review committee comprising 11 members reviewed the original English content, forward translations, synthesized version, and back translations. The committee comprised three Kiswahili language experts who participated in the forward translation and synthesis, two English language experts who conducted the back-translation, two methodological experts, two midwifery experts, one Regional Maternal and Child Health Coordinator, one Ministry of Health representative, and the Principal Investigator (PI). The committee examined linguistic accuracy, conceptual equivalence, cultural appropriateness, and clinical meaning and reached consensus on the harmonized Swahili BPCR education manual for subsequent community exploration.

#### Step 5: Community exploration

A semi-structured FGD guide was developed from the content of the harmonized Swahili BPCR manual. The guide comprised seven primary questions and 24 probes designed to explore participants’ interpretation, familiarity, semantic equivalence, cultural acceptability, and preferred terminology for BPCR concepts. The seven questions covered birth preparedness and complication readiness, danger signs during pregnancy, signs of normal labor, danger signs during labor, postpartum danger signs, newborn danger signs, and antenatal care services and attendance. The probes elicited commonly used Swahili words and expressions, more familiar alternative terms, culturally incongruent terminology or concepts, and concepts for which no familiar community expression existed. The guide was reviewed and refined before field implementation. Two research assistants received one day of training on FGD facilitation, including the use of the guide, probing techniques, management of group discussion, and ethical and confidentiality procedures. The PI also visited the selected data collection sites before field implementation to assess their suitability and prepare for the fieldwork.

### Data collection

FGDs were conducted in Swahili by the trained research assistants, who were fluent in both Swahili and Hehe. The facilitators were experienced midwives and were not from the participating health facilities, thereby reducing the potential influence of pre-existing professional relationships on participants’ responses. Discussions were conducted in private spaces suitable for group discussion. At the beginning of each FGD, the moderator explained the purpose of the study, encouraged participants to express their own views and experiences, and emphasized confidentiality. Each discussion was audio-recorded with participants’ consent, while a research assistant took field notes documenting contextual information and observations relevant to the discussion.

The harmonized Swahili BPCR manual was presented to participants during the FGDs. Participants were asked to explain the meaning of selected terms and concepts, identify expressions they commonly used, suggest more familiar alternatives where applicable, describe the cultural and social acceptability of the terminology, and explain concepts that were unfamiliar or difficult to understand. Particular attention was given to terminology related to birth preparedness, maternal danger signs, labor and delivery, postpartum danger signs, and newborn danger signs. Participants were encouraged to discuss the terms collectively and provide examples from their everyday experiences and community communication.

#### Step 6: Finalization

The findings from the community exploration were reviewed against the intended meanings of the corresponding English BPCR concepts and the clinical content of the source manual. Participant-derived terms and expressions were documented and assessed according to their linguistic, semantic, conceptual, experiential, and sociolinguistic correspondence with the intended concepts. Two linguistic experts reviewed the participant-derived expressions for linguistic and semantic equivalence, clarity, and cultural appropriateness, while two midwifery experts reviewed them for clinical accuracy and consistency with the intended maternal and newborn health concepts.

Expressions were classified as representing direct or adequate equivalence, culturally adapted equivalence, partial or limited equivalence, or no readily recognized community equivalent. Where a participant-derived expression was broader, narrower, or potentially different from the intended clinical concept, contextual clarification was incorporated to preserve the intended clinical meaning. Where no readily recognized community expression existed, the original clinical concept was retained and supplemented with explanatory descriptions, examples, demonstrations, or visual support, as appropriate. Final adaptation decisions therefore included substitution with culturally familiar terminology, contextual clarification, and supplementary explanation or visual support. The final adaptation decisions were documented in the terminology matrix, which is provided as Supplementary Table S1.

### Data processing and analysis

Audio recordings were transcribed verbatim. Because the FGDs were conducted in Swahili, the transcripts were initially maintained in the language used during data collection. English translations were prepared for analysis and reporting. The analysis followed Braun and Clarke’s six-phase thematic analysis approach. First, the research team familiarized itself with the transcripts by repeatedly reading the data and reviewing the corresponding BPCR concepts. Second, initial codes were generated from participants’ descriptions and explanations. Coding focused on semantic equivalence; conceptual equivalence, experiential expressions, cultural acceptability, sociolinguistic appropriateness, unfamiliar biomedical concepts, and requirements for additional explanation. Third, related codes were grouped into candidate categories and themes.

Fourth, candidate themes were reviewed against the original transcripts to determine whether they adequately represented participants’ accounts and whether distinct concepts had been inappropriately combined. Fifth, themes were defined and named according to their central organizing meaning. Sixth, the final thematic narrative was developed by linking participant quotations and expressions to the corresponding BPCR concepts. The analysis deliberately distinguished participant-derived findings from researcher interpretation. Participant expressions were retained as evidence, while clinical interpretation was undertaken separately to determine whether the expression adequately represented the intended BPCR concept. For culturally specific Swahili expressions, the English translation in the manuscript reflects the intended meaning rather than necessarily providing a literal word-for-word translation.

### Trustworthiness

Trustworthiness was addressed using the established qualitative criteria of credibility, dependability, confirmability, and transferability [37, 38]. Credibility was supported through the use of seven FGDs conducted across seven villages and by encouraging participants to explain, compare, and clarify terminology during group discussions. Dependability was supported through the use of a common semi-structured guide and consistent procedures for data collection, transcription, and thematic analysis. Confirmability was supported by maintaining a documented translation and adaptation process and by linking analytical themes to participant-derived expressions and quotations. Transferability was supported by describing the study setting, participants, translation process, data collection procedures, and analytical approach in sufficient detail to allow readers to judge applicability to other settings [39, 40].

### Reflexivity

The research team recognized that professional knowledge of maternal health and familiarity with biomedical terminology could influence interpretation of participants’ responses. To reduce this potential influence, the community exploration focused on participants’ own language and interpretations before clinical adaptation decisions were made. The analysis distinguished between what participants said, the meaning they attributed to a term, and the researchers’ subsequent assessment of whether the expression corresponded to the intended clinical concept [41, 42].

### Reporting guideline

The qualitative component of the study was reported in accordance with the Consolidated Criteria for Reporting Qualitative Research (COREQ), including reporting of the research team and reflexivity, participant selection, data collection procedures, data analysis, and presentation of participant quotations [43].

### Ethics approval, consent, and confidentiality

As the study involved human participants, ethical clearance and a permission letter were obtained from the Research Review Committee of the Research and Publication Office at the University of Dodoma prior to data collection (number MA.84/261/86/11). A written informed consent was obtained from all participants before their inclusion in the study. All participants were above 18 years. All procedures performed in this study involving human participants were conducted in accordance with institutional ethical standards and principles. Permission to conduct the study in the Iringa region was obtained from the Permanent Secretary, President’s Office, Regional Administration, and local government reference number AB.307/323/01“R”/70. Permission to use the ANC guidelines for the study was requested from the Ministry of Health, reference number PA.104/282/02. The permission to collect data in the Iringa region was obtained from the Iringa Regional Secretary, reference number Na.FA.225/265/01/J/281. The permission to collect data in the Mufindi and Kilolo districts was obtained from district directors; in the Mufindi district, the permission reference number was Na. HW/MUF/S.40/43/VOL.IV/39, and the Kilolo reference number was Na. KDC/S.10/4/VOLL.VIII/193. Prior to data collection, written informed consent was obtained from all participants. Throughout the study, participant confidentiality and privacy were strictly maintained by assigning unique identification numbers instead of using personal names. Cultural respect and sensitivity were carefully observed during all stages of the study to ensure that the training content, delivery, and implementation were culturally appropriate and did not conflict with participants’ beliefs, values, or customary practices.

## Results

### Participant characteristics

A total of 56 pregnant women participated in seven FGDs conducted across seven villages in Kilolo and Mufindi districts. Participants had a mean age of approximately 28.4 years. Most had previous pregnancy and childbirth experience, had completed primary education, and were married. The seven FGDs generated detailed accounts of how participants understood, interpreted, and expressed terminology contained in the harmonized Swahili BPCR education manual.

### Overview of thematic findings

Five themes were developed:

Theme 1: Culturally and linguistically familiar expressions conveyed BPCR concepts.

Theme 2: Experiential and contextual language shaped descriptions of danger signs.

Theme 3: Sociolinguistic norms and modesty influenced communication about sensitive health topics.

Theme 4: Some clinically important concepts had partial or limited conceptual equivalence.

Theme 5: Unfamiliar concepts required supplementary explanation.

The findings demonstrated that equivalence varied across BPCR concepts. Some concepts had familiar expressions that conveyed the intended meaning, some required contextual clarification, and others lacked a readily recognized community equivalent.

### Theme 1: Culturally and linguistically familiar expressions conveyed BPCR concepts

Participants generally recognized the major BPCR concepts presented in the harmonized Swahili manual. However, they identified expressions that they considered more natural and meaningful in everyday community communication.

#### Anticipated date of delivery

Participants distinguished between *“matarajio ya kujifungua”* and *“matazamio ya kujifungua.”* Participants perceived the former as suggesting greater certainty about the timing of delivery, whereas the latter was understood as expressing anticipation without implying that delivery would necessarily occur on a specific date.

One participant explained: “We say ‘anticipated date of delivery’ rather than ‘expectations,’ because when you say ‘expectations,’ you are certain that this thing must happen. But when you say ‘anticipating,’ it means it could happen either before or after the due date.” (P36). The participant’s explanation illustrates that apparently similar terms could carry different meanings regarding certainty and timing.

#### Family support and birth preparedness

Participants described preparation for childbirth using expressions that reflected family roles. For example, *“kuandaa ndugu wa kumsindikiza mama mjamzito kituoni”* referred to preparing a relative to accompany a pregnant woman to the health facility. The expression *“mtu wa kukaa na familia nyumbani”* referred to identifying someone who would remain at home to care for family members while the pregnant woman was away for delivery. These expressions were used to describe practical arrangements within the family rather than abstract concepts of birth preparedness.

#### Emergency transport and financial preparation

Participants used the expression *“fedha ndiyo usafiri”* (“money itself is transport”) when discussing preparation for emergency transportation. The expression connected the availability of money with the ability to obtain transport when an emergency occurred. Participants therefore framed transport preparedness not only as identifying a vehicle but also as ensuring the financial resources necessary to access it. The expression was treated as a culturally meaningful representation of emergency transport preparation while retaining the underlying clinical message.

### Theme 2: Experiential and contextual language shaped descriptions of danger signs

Participants frequently described danger signs through observable experiences and bodily changes rather than formal biomedical terminology.

#### Rupture of membranes

Participants used *“chupa imepasuka”* (“the water bag has burst”) and *“chupa kupasuka mapema”* (“the water bag has burst early”) to describe rupture of membranes. One participant stated: “We say *chupa imepasuka*.” (P5). The expression was familiar to participants and referred to the experience of the membranes breaking before or during labor. The term was therefore retained as a culturally familiar expression while the clinical meaning was maintained in the adapted material.

#### Absent fetal movement

Participants used *“mtoto kutokucheza tumboni”* (“the baby is not moving in the womb”) to describe absent or markedly reduced fetal movement. One participant explained: “We say the baby is not moving at all in the womb.” (P51). The expression describes an observable change in fetal movement and reflects the way participants described the experience in everyday language.

#### Delayed or prolonged labor

Participants used *“kuchelewa kujifungua”* (“delay in giving birth”) when describing prolonged labor. One participant stated: “We say the woman has delayed in giving birth.” (P29). The expression was broader than the biomedical definition of prolonged labor because it did not specify a particular duration. It was therefore treated as a participant-derived description rather than a direct substitute for the clinical criterion.

#### Other experiential expressions

Participants used *“mtoto kukaa vibaya tumboni”* (“the baby is positioned wrongly in the womb”) to describe fetal malpresentation. For newborn fever, participants used expressions such as *“mtoto kuchemka”* or *“amechemka”*, referring to a newborn who felt very hot. Participants also described inability to pass stool and urine using *“mtoto hapati choo na mkojo”* (“the baby does not pass stool and urine”). These expressions were documented as examples of experiential terminology and reviewed against the corresponding clinical concepts.

### Theme 3: Sociolinguistic norms and modesty influenced communication about sensitive health topics

Participants described social expectations concerning how reproductive anatomy and other sensitive maternal health topics should be discussed.

#### Reproductive anatomy

Participants used *“sehemu za siri”* (“private parts”) rather than the direct anatomical term *“uke”* (“vagina”) in some communication contexts. One participant explained: “We use *‘sehemu za siri’* (‘private parts’) to soften the harshness of words, because in the Hehe community, mentioning ‘ukeni’ is considered a serious insult.” (P53). Another participant stated: “Even though people understand what ‘uke’ means, as my colleague explained, I cannot, for example, go to my mother-in-law and say, ‘Mother, I am bleeding from my vagina.’ She would understand, but she would perceive it as using an offensive word. That is why the term *‘sehemu za siri’* is used.” (P54).

These accounts demonstrate that understanding and social acceptability were distinct considerations. Participants could understand a direct anatomical term while nevertheless preferring a euphemistic expression in certain interpersonal settings.

#### Fetal position and other sensitive descriptions

Participants used expressions such as *“mtoto kukaa vibaya tumboni”* (“the baby is positioned wrongly in the womb”) when discussing fetal malpresentation. The expression was socially familiar and experiential, but it did not specify the particular type of fetal malpresentation. Consequently, clinical clarification remained necessary.

### Theme 4: Some clinically important concepts had partial or limited conceptual equivalence

The discussions identified several concepts for which participants had limited familiarity or where the community expression was broader than the biomedical concept.

#### Cord prolapses

Participants did not identify a readily recognized community expression for cord prolapse. Some reported that they had never seen or heard about the umbilical cord coming out before the baby.

Participants stated: “We do not know whether the umbilical cord comes out first, and we have never seen nor heard about it.” (P34–41; P42–50; P51–56). The response indicated limited familiarity with the clinical event rather than merely a lack of vocabulary. The concept was therefore not replaced with an arbitrary local expression. Instead, additional explanation was considered necessary.

#### Neonatal cyanosis

Participants also reported limited familiarity with neonatal cyanosis, particularly bluish discoloration of the lips, tongue, or hands. One participant stated: “We have never seen it happen to a baby, and we have never seen any baby with blue color.” (P15). Another stated: “I cannot answer because I have never seen a baby whose lips, tongue, and hands are blue.” (P29). Participants contrasted this unfamiliarity with neonatal jaundice, which they had encountered more often. One participant stated, “We have never seen a blue-colored baby or a baby that changes color, but we have seen a yellow-colored baby.” (P16). The findings indicate that the challenge was conceptual and experiential as well as linguistic.

#### Prematurity and low birth weight

Participants used *“mtoto kuzaliwa njiti”* (“a baby born prematurely”) when discussing premature birth. However, the participant-derived expression did not distinguish prematurity from low birth weight. Because prematurity and low birth weight are related but clinically distinct concepts, the terminology required clarification rather than being treated as interchangeable.

#### Partial equivalence

Other expressions, including *“kuchelewa kujifungua”* for prolonged labor and *“mtoto kukaa vibaya tumboni”* for fetal malpresentation, were familiar to participants but did not contain all the information represented by the corresponding biomedical terminology. These examples were therefore classified as partial or experiential equivalence.

### Theme 5: Unfamiliar concepts required supplementary explanation

Participants’ responses showed that some concepts could not be adequately communicated through a translated term alone.

#### Need for explanatory descriptions

For cord prolapse and neonatal cyanosis, participants lacked a readily recognized local expression or experiential reference. In such circumstances, replacing a biomedical term with another word would not necessarily resolve the communication problem. Participants’ responses suggested the need for explanations based on observable features and contextual examples.

#### Need for visual or demonstrative support

The difficulty participants experienced in describing neonatal cyanosis illustrated the potential value of visual support. Participants could recognize neonatal jaundice but reported that they had not encountered a blue-colored newborn. The findings therefore indicated that visual or demonstrative approaches may be needed for concepts that were difficult to communicate using locally familiar terminology. The same principle applied to cord prolapse, for which the concept itself was unfamiliar to participants.

#### Summary of cultural and linguistic equivalence patterns

The community exploration identified four broad patterns in how participants interpreted and expressed the translated BPCR terminology:

**1. Direct or adequate equivalence:** Translated terminology was readily understood and corresponded closely with expressions familiar to participants.
**2. Culturally familiar equivalence:** Participants identified commonly used community expressions that conveyed the intended BPCR concept in a culturally familiar manner.
**3. Partial equivalence:** a familiar community expression was identified, but its meaning was broader, narrower, or different from the intended biomedical concept and required further explanation or contextual interpretation.
**4. No readily recognized equivalent:** Participants could not identify a familiar community expression or experiential reference point for some concepts, indicating areas where the translated terminology was not readily recognizable within the community.

These patterns illustrate the range of cultural and linguistic equivalence identified during community exploration and provide the basis for reporting the culturally familiar BPCR terminology identified by participants. The complete terminology matrix is provided in Supplementary Table S1.

## Discussion

### Principal findings

This study explored how Hehe pregnant women interpreted BPCR terminology contained in a harmonized Swahili education manual. The findings demonstrate that cultural and linguistic equivalence was not uniform across concepts. Participants frequently used expressions grounded in everyday experience, family relationships, bodily observations, and culturally acceptable communication practices. At the same time, several clinical concepts were unfamiliar or lacked a readily recognized community expression.

The findings therefore show that translation and cultural adaptation should be understood as related but distinct processes. Translation establishes linguistic meaning, whereas cultural exploration determines whether that meaning corresponds with how members of the target population understand and communicate the concept [44, 45].

### Beyond word-for-word translation

The distinction participants made between *“matarajio ya kujifungua”* and *“matazamio ya kujifungua”* illustrates how apparently similar expressions can carry different meanings. Participants associated *matarajio* with greater certainty, whereas *matazamio* was understood as allowing for variation in timing. This finding supports the principle of dynamic equivalence, which emphasizes communication of intended meaning rather than reproduction of source-language wording [29]. In health education, such distinctions can be particularly important when terminology communicates probability, timing, severity, or urgency [46, 47].

The implication is not that one linguistic expression is universally superior to another. Rather, the findings demonstrate the importance of testing translated terminology with intended users before finalizing health education materials [48, 49].

### Experiential language and maternal danger signs

Participants commonly described danger signs through observable experiences. Expressions such as *“chupa imepasuka”* and *“mtoto kutokucheza tumboni”* provide examples of terminology grounded in experiences that pregnant women can recognize. The use of experiential descriptions is important because many biomedical terms are abstract and may not correspond directly to how community members describe bodily changes. Similar research on maternal health communication has shown that recognition of pregnancy complications can be influenced by women’s experiences, local explanatory models, and community terminology [50, 51, 52].

However, the findings also demonstrate the need to preserve clinical precision. For example, *“kuchelewa kujifungua”* communicates delayed childbirth but does not itself define the clinical duration of prolonged labor. Likewise, *“mtoto kukaa vibaya tumboni”* conveys abnormal fetal positioning but does not specify the particular clinical presentation. Cultural adaptation should therefore not simply replace biomedical terminology. Instead, community expressions should be used when they communicate the intended concept, with clinical clarification added where necessary [53, 54].

### Sociolinguistic acceptability and reproductive terminology

The preference for *“sehemu za siri”* demonstrates that cultural equivalence includes sociolinguistic acceptability. Participants could understand direct anatomical terminology but considered it inappropriate in some social relationships. This distinction between comprehension and acceptability is important for maternal health education. A technically accurate term may be less suitable for community communication if it violates locally recognized norms of politeness or modesty [55, 56].

At the same time, euphemistic terminology should not be allowed to introduce ambiguity. The appropriate approach is therefore to use culturally acceptable language while maintaining enough clinical explanation to ensure that the intended meaning is clear [57, 58].

### Limits of Cultural Equivalence

The findings concerning cord prolapse and neonatal cyanosis demonstrate the limits of linguistic adaptation. Participants did not identify a familiar expression for cord prolapse and reported little experience with the condition. Similarly, participants were unfamiliar with neonatal cyanosis and contrasted the unfamiliar concept of a “blue baby” with the more familiar “yellow baby.” These findings show that not every biomedical concept has a culturally equivalent community term. Attempting to create a local expression simply to achieve linguistic equivalence could produce terminology that is inaccurate or confusing [59, 60, 61]. For such concepts, adaptation should instead focus on conceptual explanation. Clear descriptions of observable features, examples, demonstrations, and visual materials may be more appropriate than substitution of terminology alone [62].

The findings also showed that participant-derived expressions could be meaningful and culturally familiar while remaining broader than the corresponding clinical concepts. For example, *“mtoto kuzaliwa njiti”* refers to a baby being born prematurely but should not be treated as synonymous with low birth weight [63]. Similarly, *“kuchelewa kujifungua”* communicates delayed childbirth but does not necessarily correspond to the biomedical definition of prolonged labor [64]. These examples highlight an important limitation of cultural equivalence: cultural familiarity does not necessarily imply clinical equivalence [53, 65]. Therefore, culturally familiar expressions should be evaluated in relation to the intended clinical meaning to ensure that adaptation preserves both comprehensibility and clinical accuracy [66, 67].

### Contribution to the literature

The main contribution of this study is its demonstration of a community-based approach to identifying cultural and linguistic equivalence within translated maternal health education content. The study contributes in three ways. First, it demonstrates that target-community exploration can identify distinctions that may not be apparent during professional translation alone. Second, it provides empirical examples of how Hehe pregnant women describe BPCR concepts using everyday Swahili expressions. Third, it identifies a practical distinction among direct equivalence, culturally adapted equivalence, partial equivalence, and concepts requiring supplementary explanation. This framework may be useful for other maternal health education materials translated for linguistically and culturally diverse populations. The study also contributes evidence from the Hehe community, which is comparatively underrepresented in research on cultural adaptation of maternal health education.

### Implications for development of BPCR education materials

The findings support the involvement of intended users in the development and refinement of translated health education materials. The translation process can be strengthened through a sequential approach that includes translating the source material, reviewing translation discrepancies, conducting expert review, exploring terminology with members of the target population, distinguishing culturally familiar expressions from clinically equivalent expressions, and adding explanatory or visual support where no adequate community equivalent exists. This approach allows culturally meaningful expressions to be incorporated without assuming that all community expressions are automatically clinically equivalent. These findings provide an empirical basis for the subsequent refinement of BPCR educational terminology to improve its cultural and linguistic appropriateness.

### Strengths and limitations

#### Strengths

This study had several strengths. First, it directly involved intended users of the BPCR education material in the cultural and linguistic exploration process. Second, the seven FGDs were conducted across seven villages in two rural districts, allowing terminology to be explored across multiple community settings. Third, the use of FGDs facilitated interaction among participants and enabled participants to compare and explain alternative expressions. Fourth, community exploration was preceded by a structured translation, synthesis, back-translation, and expert-review process. Fifth, the analysis explicitly distinguished between linguistic familiarity, cultural acceptability, experiential equivalence, and clinical meaning.

#### Limitations

The study also had limitations. First, it was conducted among Hehe pregnant women in two rural districts of the Iringa Region. The findings may therefore not be directly transferable to other ethnic, linguistic, or geographical populations. Second, FGDs are subject to group dynamics and social desirability. Participants may have been influenced by the views of other participants or may have avoided terminology considered socially sensitive. Third, participants’ interpretations were explored qualitatively rather than tested through a formal comprehension assessment. Therefore, the study cannot establish quantitatively that the adapted expressions produce greater comprehension than the original translated terminology. Fourth, some clinical concepts were unfamiliar to participants. The absence of a community expression should not be interpreted as evidence that the condition is absent from the population. Fifth, some participant-derived expressions were broader than their biomedical counterparts. This created a continuing need for clinical review during adaptation. Finally, because the study focused on terminology and conceptual interpretation, it did not evaluate the effect of the adapted manual on BPCR knowledge, attitudes, practices, care-seeking behavioural, or maternal and newborn health outcomes.

## Conclusion

This study found that cultural and linguistic equivalence of BPCR education content involved more than translating English biomedical terminology into Swahili. Hehe pregnant women frequently used culturally familiar and experiential expressions to describe BPCR concepts, including *“matazamio ya kujifungua,” “fedha ndiyo usafiri,” “chupa imepasuka,” “mtoto kutokucheza tumboni,”* and *“sehemu za siri.”* While some expressions conveyed the intended clinical concepts adequately, others were broader than their biomedical equivalents and required clarification. Other concepts, particularly cord prolapse and neonatal cyanosis, lacked readily recognized community equivalents and therefore required alternative explanatory approaches.

The findings demonstrate the value of involving intended users in the cultural and linguistic adaptation of maternal health education materials. Community-derived terminology can contribute to culturally meaningful communication when it is evaluated against the intended clinical meaning. Where no adequate community equivalent exists, explanatory descriptions and visual support may be preferable to creating an artificial translation.

However, this exploratory study does not establish the effectiveness of the adapted terminology or educational manual. Its effects on BPCR knowledge, attitudes, practices, health-seeking behaviors, and maternal and newborn outcomes require evaluation in appropriately designed intervention studies.

## Abbreviations

ANC: Antenatal Care
BPCR: Birth Preparedness and Complication Readiness
FGD: Focus Group Discussion

## Consent for publication

Not applicable.

## Availability of data and materials

Because the qualitative data contain information obtained from individual participants, the full transcripts are not publicly available. Selected participant quotations supporting the findings are presented in the manuscript. The complete cultural and linguistic terminology matrix is provided as supplementary material.

## Data Availability

The qualitative data generated during this study are not publicly available because they contain potentially identifiable information and sensitive participant narratives. Data may be made available from the corresponding author upon reasonable request, subject to ethical approval and applicable data-protection requirements.

## Acknowledgements

The authors thank the pregnant women who participated in the focus group discussions and the health-facility teams and community members who supported the study. The authors also acknowledge the linguistic and technical experts who contributed to translation, review, and refinement of the BPCR education content.

## Authors’ contributions

RBM conceived and designed the study, coordinated the translation and cultural adaptation process, contributed to data collection and analysis, and drafted the manuscript. SAS, ROS, and FVM contributed to the study design, methodological review, interpretation of findings, and critical revision of the manuscript. All authors reviewed and approved the final manuscript.

## Supplementary material

**Supplementary Table S1.**
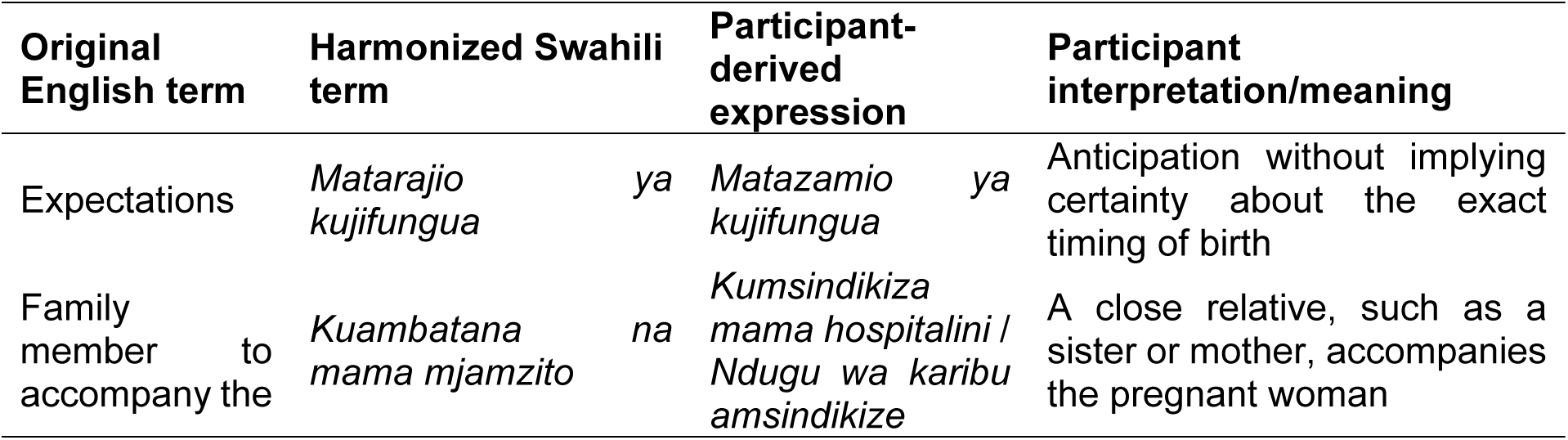
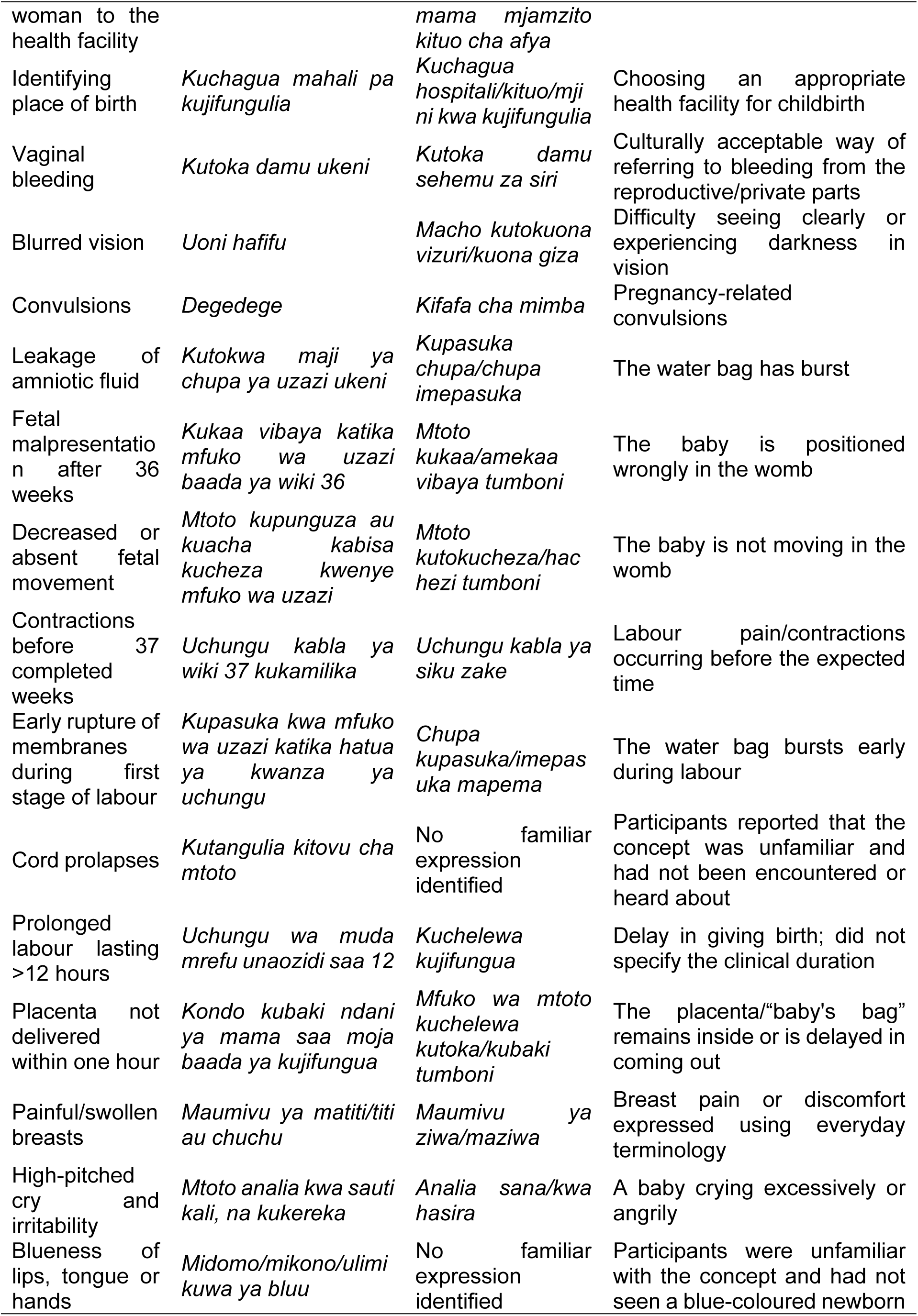
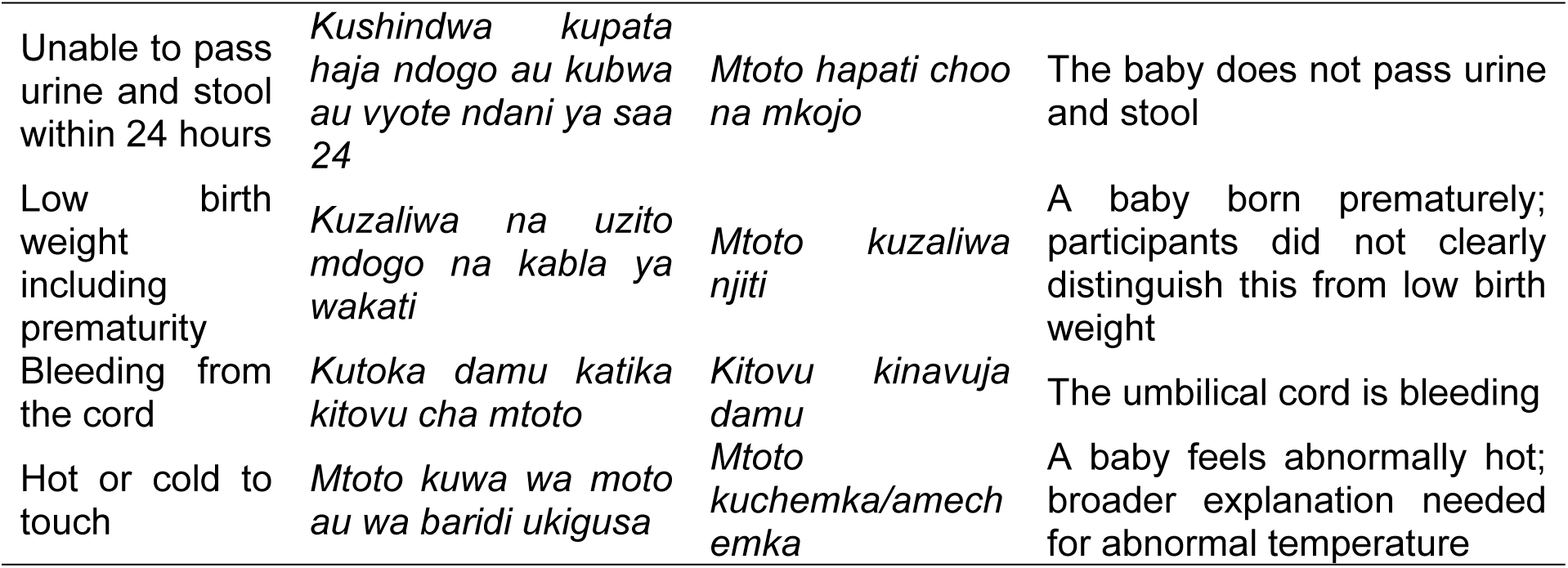
Complete Cultural and Linguistic Equivalence Matrix for BPCR Terminology.

